# Pembrolizumab alters the tumor immune landscape in a patient with dMMR glioblastoma

**DOI:** 10.1101/2023.12.08.23299732

**Authors:** Todd Bartkowiak, Asa A. Brockman, Bret C. Mobley, Hannah Harmsen, Paul Moots, Ryan Merrell, Douglas B. Johnson, Reid C. Thompson, Vinay K. Puduvalli, Rebecca A. Ihrie

## Abstract

Congenital DNA mismatch repair defects (dMMR), such as Lynch Syndrome, predispose patients to a variety of cancers and account for approximately 1% of glioblastoma cases. While few therapeutic options exist for glioblastoma, checkpoint blockade therapy has proven effective in dMMR tumors. Here we present a case study of a male in their 30s diagnosed with dMMR glioblastoma treated with pembrolizumab who experienced a partial response to therapy. Using a multiplex IHC analysis pipeline on archived slide specimens from tumor resections at diagnosis and after therapeutic interventions, we quantified changes in the frequency and spatial distribution of key cell populations in the tumor tissue. Notably, proliferating (KI67+) macrophages and T cells increased in frequency as did other KI67+ cells within the tumor. Therapeutic intervention remodeled the cellular spatial distribution in the tumor leading to a greater frequency of macrophage/tumor cell interactions and T cell/T cell interactions, highlighting impacts of checkpoint blockade on tumor cytoarchitecture and revealing spatial patterns that may indicate advantageous immune interactions in glioma and other solid tumors treated with these agents.

**Insight:** This work sheds light on the capacity of checkpoint blockade therapy to modulate the immune microenvironment in DNA mismatch repair deficient glioblastoma, highlights the utility of window-of-opportunity clinical trials in patient selection of immunomodulatory therapies, and demonstrates the feasibility and utility of mapping cellular interactions associated with therapeutic responses in gliomas and other solid tumors.

**Statement regarding non-clinical trial status:** We confirmed with a treating neurologist in this case that the treatment received by the individual whose samples are studied was part of routine clinical care and not a clinical trial, as the patient was previously diagnosed with Lynch syndrome (a mismatch repair deficiency). The use of pembrolizumab is recommended for treatment of tumors with high mutational burden due to mismatch repair deficiency and is currently considered standard of care for these tumors. Additionally, though the intervention and outcome are detailed in the manuscript, the focus of the manuscript is on reporting changes observed in the immune microenvironment at different points in the clinical trajectory - a retrospective analysis performed after clinical care was complete.

## Introduction

Lynch Syndrome is an autosomal dominant genetic condition whereby mutations in the DNA mismatch repair (dMMR) machinery increase the risk of cancer development including colorectal, endometrial, gastric, and ovarian cancers (1). The most common Lynch Syndrome-associated mutations occur in the *MutL-like homolog 1* (*MLH1*) (20-40% of cases), *MutS homolog 2* (*MSH2*) (25-60%), or *MSH6* (10-40%) tumor suppressor genes. These mutations, however, are only found in 1% of glioblastoma cases overall and therapeutic options for dMMR glioblastomas are limited. Basket trials have proven checkpoint blockade therapy (CPB) efficacy in dMMR/MSI-high colorectal carcinoma, suggesting a comparable feasibility for brain cancers (2). A detailed understanding of the immune infiltrate – both its composition and cytoarchitecture – provides a valuable opportunity to identify cellular interactions associated with therapeutic response.

Advances in multiplex imaging analyses have highlighted the capacity of cellular topography within tumors to help identify spatial patterns correlated with patient outcomes and response to therapy (3-5). Here 10-round cyclic immunohistochemistry was used to quantify the abundance of 24 cell types within the tumor tissue of a patient’s dMMR glioma throughout the course of therapy, identify the spatial relationships between cells, and illuminate changes in these features in response to CPB (**Supplementary Methods**). Therapeutic intervention with pembrolizumab modulated the frequency and distribution of myeloid and lymphoid components within resected primary and recurrent dMMR tumor samples. This work highlights potential combinatorial therapeutic strategies for dMMR-associated glioblastoma, and the potential for IHC to assess changes in the tumor-immune microenvironment as a secondary clinical endpoint during clinical trials including window-of-opportunity trials.

### Case Presentation

A male in their early 30s previously diagnosed with Lynch Syndrome and a positive family history presented with near syncope, intermittent left-side head pain lasting 30-60 minutes, aphasia, and blurred vision with no other seizure-like symptoms in the previous four months. The patient was placed on indomethacin. MRI imaging showed a left temporal lobe enhancing mass with hemorrhaging, surrounding edema and brain stem mass effect. The patient was diagnosed with an *IDH*-WT, uMGMT glioblastoma and received maximum safe surgical resection with concurrent radiation (60Gy) and temozolomide (TMZ). Next generation sequencing revealed a pathogenic LOF mutation in *MLH1* (c.380+1G>A) and a tumor mutation burden of 19.5 mutations/MB confirming a diagnosis of Lynch Syndrome. After four months, the patient was placed on monthly temozolomide and Tumor-Treating Field (TTF) therapy (Optune). 11 months later, the patient was placed on pembrolizumab (200 mg Q3W). The patient’s tumor was biopsied after two months, and he received intraoperative Laser Interstitial Thermal Therapy (LITT) with continued pembrolizumab for an additional 3 months before receiving another biopsy with confirmed active tumor. The patient was then placed on bevacizumab and lomustine (CCNU).

## Results

### Immune and tumor stromal abundance is impacted by therapeutic intervention

To determine the frequency and spatial distribution of immune and tumor cells in dMMR glioblastoma, tumor tissue sections were collected 1) at time of diagnosis, 2) at first recurrence after sequential radiation/TMZ therapy, TMZ/TTF, and single-agent pembrolizumab (200 mg/kg Q3W) and 3) single-agent pembrolizumab after intraoperative LITT therapy (**Figure 1A**). Ten rounds of cyclic immunohistochemistry assessed eight immune targets (**Supplementary Methods**, **Supplementary Table 1, Figure 1B**). Each round of images was collected, aligned, and CD3+ or CD68+ immune and CD3-CD68-populations (here, subsequently termed “tumor” for simplicity) were expertly gated (**Supplementary Figure 1 A-D, Supplementary Figure 2**)(6). Changes in the frequency of lymphoid and myeloid populations were found throughout the course of therapy (**Figure 1C-E**, **Supplementary Figure 3**). Notably, microglia were absent in the tumor microenvironment after pembrolizumab following TMZ/TTF (resection 2) and the macrophage compartment shifted from a PD-L1^-^KI67^-^ to a PD-L1^+^ phenotype. A population of PD-L1^+^KI67^+^ macrophages appeared after CPB in the secondary and tertiary resection while the frequency and density of PD-L1^-^K67^-^ and PD-L1^+^ macrophages decreased. Although lymphocyte populations were depleted in the secondary resection, the proportion and density of PD-1^-^KI67^-^ lymphocytes increased subsequent to pembrolizumab after LITT therapy (resection 3).

**Figure 1:**
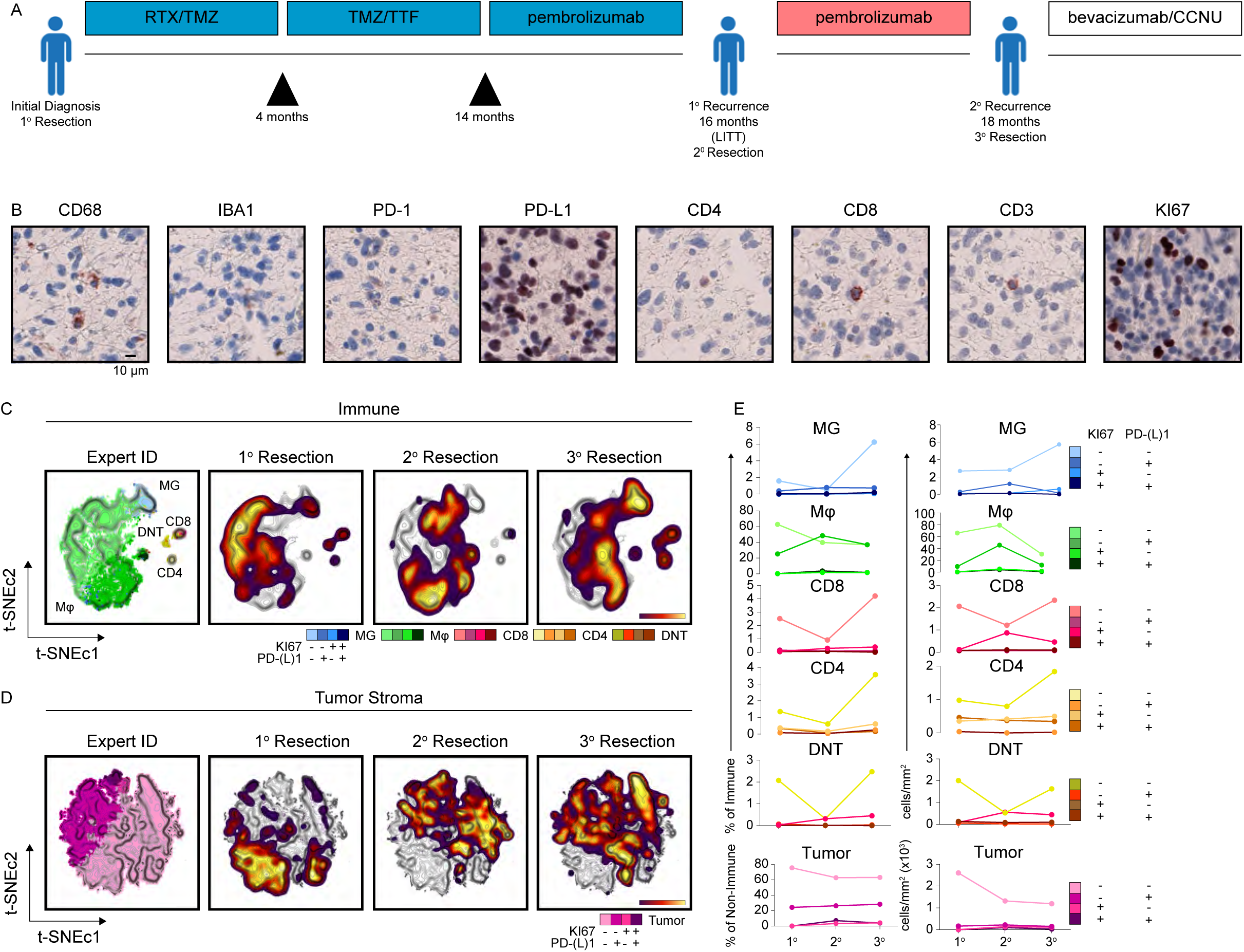
Altered immune and tumor cell frequencies throughout the course of therapeutic intervention. A) Schema indicating the patient’s treatment strategy. B) Representative IHC micrographs for each antibody marker assessed. C) t-SNE-CUDA analysis of the CD3^+^CD68^-^ or CD3^-^CD68^+^ immune fraction of each tumor sample. Manually gated populations were overlaid onto the t-SNE map of cells pooled from each sample to indicate the relative positions of indicated immune populations on the map. Density contours indicate the cellular fraction from the primary, secondary, or tertiary resection overlaid onto the contour. D) t-SNE-CUDA analysis of the CD3^-^ CD68^-^ fraction of the tissue. E) Graphs indicating the frequency (left) or cell density (right) of each indicated subset of microglia (MG; blue), macrophages (Mφ; green), cytotoxic T cells (CD8; red), helper T cells (CD4; yellow) or CD8^-^CD4^-^ cells (DNT; orange) as a percent of the scored immune fraction in the tissue or of tumor (Tumor; pink) as a percent of CD3^-^CD68^-^ cells. Scale bar in B indicates 10 µm. RTX = radiation, TMZ = temozolomide, TTF = Tumor Treating Fields, LITT = Laser Interstitial Thermal Therapy, CCNU = 1-(2-chloroetyl)-3-cyclohexyl-1-nitrosurea (lomustine).

Assessment of CD3^-^CD68^-^ tumor cells during the course of therapy revealed similar findings as the immune fraction (**Figure 1D**). The first resection was largely comprised of PD-L1^-^ KI67^-^ tumor cells and a smaller fraction of PD-L1^+^KI67^-^ cells (**Figure 1E**, **Supplementary Figure 3B**). After treatment with pembrolizumab (resections 2 and 3), PD-L1^-^KI67^+^, PD-L1^+^KI67^-^, and PD-L1^+^KI67^+^ cells were more prevalent in the tissue.

Taken together, these data suggest that CPB alters the frequency and phenotype of immune and other stromal cells in dMMR glioblastoma. Interestingly, therapy had a considerable impact on cellular proliferation, as KI67+ tumor and phagocyte populations increased throughout the course of therapy. Secondly, therapy dramatically remodeled the phagocytic pool in the tumor by reducing microglia and increasing PD-L1^+^ macrophages.

### Altered spatial distribution of immune and tumor stromal cells during therapy

Next, the impact of CPB on the spatial relationships between immune and tumor cells was assessed (**Figure 2A**). Mapping the expert-gated cell populations back to their positions within the tissue (**Figure 2A-B**, **Supplementary Figure 4**), the composition, frequency, and location of the 15 most abundant cellular neighborhoods in the tissue were scored (**Figure 2C**, **Supplementary Figure 5A-F, Supplementary Figure 6).** These data were used to identify non-random cellular interactions that may correlate with biological function (as in known structures such as germinal centers in tonsil, Neighborhood 15, **Supplementary Figure 5G**).

**Figure 2:**
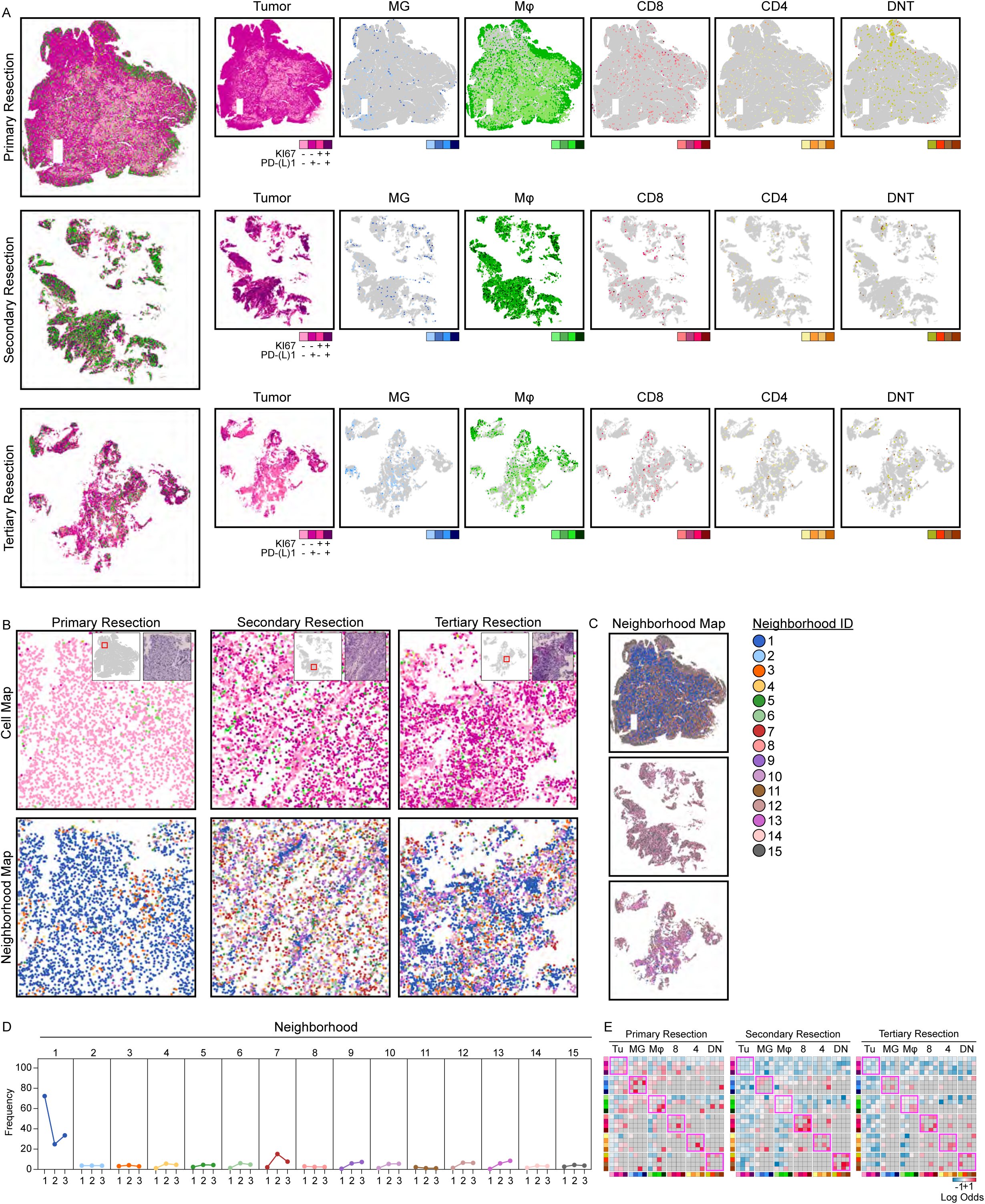
Altered neighborhood distribution in tumor tissue throughout the course of therapy. A) Expert-gated populations mapped to their original locations within the tumor tissue in the primary (top), secondary (middle), or tertiary (bottom) resection. B) Representative regions-of-interest within each resected tumor tissue indicating the position of expert-gated populations (top) or cell neighborhood identity (bottom). C) Tissue map indicating the spatial position of each cellular neighborhood across each tumor resection. D) Frequency of each cellular neighborhood within each resected tissue sample. E) Log-odds ratio clustergrams indicating the odds ratio of each potential cell-cell interaction occurring in the primary (left), secondary (middle), or tertiary (right) resected tumor tissue. Insets in B indicate the location of the region-of-interest (red box) within the tissue and the H&E staining of the ROI. Log-odds ratio plots in E indicate the likelihood of two cellular interactions occurring more frequently (LOR>1, red) or less frequently (LOR<-1, blue) than expected given each cell types frequency distribution in the tissue.

The spatial distribution of immune and tumor cells was altered in post-treatment samples (**Figure 2B, C)** as were the position and frequency of cellular neighborhoods in the tissue (**Figure 2D-E; Supplementary Figure 6, Supplementary Figure 7**). The majority of the pretreatment tissue (resection 1) consisted of neighborhood 1, composed entirely of PD-L1^-^KI67^-^ cells (**Figure 1D, Supplementary Figure 6, Supplementary Figure 7A**) whereas neighborhoods 2, 5, and 7 containing PD-L1+ tumor cells appeared near tissue edges. Intriguingly, not only did the frequency of neighborhood 1 (PD-L1^-^KI67^-^ stroma) decrease after therapy, but the frequency of neighborhood 7 (PD-L1+ and PD-L1-macrophages proximal to PD-L1+ and KI67+ tumor) (**Supplementary Figure 6**) increased after pembrolizumab, as did neighborhoods 9, 10, 12, and 13 (**Figure 2B-D**). Moreover, the tissue distribution of cellular neighborhoods was affected by checkpoint blockade therapy. For instance, neighborhoods 2, 4, 5, 6, 7, 10, 12, 14, and 15 presented along the edge of the primary resection but were distributed throughout the secondary and tertiary resection (**Supplementary Figure 7 A-C**).

In addition to identifying the most common cellular neighborhoods, the log-odds ratio was used to identify statistically enriched cellular interactions while accounting for the frequency of each cell type (**Supplementary Figure 5G**) (7). Using this approach, Ki67+ macrophage/macrophage interactions occurred more frequently in the primary resection, while CD8 T/CD8 T cell interactions occurred more frequently after TTF/CPB (resection 2) (**Figure 2E**). Mapping the log-odds ratio of cell-cell interactions across the entire tissue identified regions where specific cell-cell interactions occurred (**Supplementary Figure 8, Supplementary Figure 9**). For instance, germinal centers (orange tiles) were identified in the positive control sample, tonsil (**Supplementary Figure 7F, Supplementary Figure 8**). Within the GBM tissue sections, regions with greater T cell interactions were found in tumor samples after treatment with pembrolizumab, (orange and blue boxes in Supplementary Figure 5 B-C) which were prevalent after LITT therapy in resection 3.

These data quantitatively reveal how immunotherapeutic intervention in combination with either TTF or LITT therapy modulated the immune and tumor stromal components within the glioma microenvironment. Particularly, regions with abundant macrophage/tumor and/or T cell/T cell interactions may be conducive to therapeutic efficacy particularly when CPB is administered.

## Conclusion

Glioblastoma remains an incurable tumor despite aggressive treatment; however tumor-lysate-loaded dendritic cell vaccines in newly diagnosed or recurrent GBM highlight the potential for effective immunotherapeutics (8). The complexities that mediate resistance to immunotherapy in GBM remain unresolved. One way to improve the clinical utility of immunotherapy for resistant tumors like GBM may lie in selecting patients with dMMR tumors or using genotoxic neoadjuvant therapies that increase immunostimulatory neoantigen expression. PD-1-targeted therapy reduced tumor burden and extended survival in MSI-high/dMMR cancers (2), warranting further exploration in brain tumors. This study highlights the capacity for CPB to modulate the immune microenvironment in a patient with dMMR glioblastoma. LITT and pembrolizumab increased the abundance of CD8, CD4, and DNT cells. TTF therapy depleted microglia in the tumor and increased PD-L1^+^KI67^+^ macrophages. In addition to inducing tumor apoptosis, TTF may polarize macrophages towards an inflammatory phenotype (9). Indeed, PD-L1 expression indicates an inflammatory milieu that may respond to immunotherapy. Moreover, intraoperative LITT may complement immunotherapy by increasing antigen release (10). Interestingly, post-therapy samples showed evidence of increased proliferation in macrophages and tumor cells. While macrophages are commonly associated with therapeutic resistance, it remains unclear whether the macrophages in this patient were pro- or anti-inflammatory. Moreover, future studies will need to determine whether therapeutic intervention in this patient led to expansion of aggressive proliferating tumor cells, or if the CD3-CD68-population included inflammatory immune components (e.g. B cells, NK cells, or dendritic cells). This work highlights the potential for Window-of-Opportunity (WOO) clinical trial design for glioma patients whereby resected tissue is collected for analysis prior to and/or during therapy and changes in the immune and tumor composition can be assessed in conjunction with clinical data.

## Data Availability

Datasets analyzed in this manuscript will be made available for reviewers and at the time of publication. Transparent analysis scripts for datasets in this manuscript are publicly available on the Ihrie Lab Github page (https://github.com/ihrie-lab/CASSATT) with open-source code and commented python analysis walkthroughs.

## Acknowledgements

We thank the Vanderbilt Translational Pathology Shared Resource, the Vanderbilt Neurovisualization Laboratory, and the surgeons, patient, and family that supported this work.

## Author Contributions

T.B. designed the study. T.B., R.A.I., and P.M. conceptualized the study. T.B. and A.A.B collected data. A.A.B developed the image analysis workflow. T.B. and A.A.B. analyzed data. P.M., R.M., D.B.J., R.C.T., and V.K.P. provided patient care. B.C.M., P.M., V.K.P. and H.H. provided patient tissue samples and clinical information. T.B. and R.A.I. wrote the manuscript. R.A.I. provided financial support. All authors contributed to reviewing and editing the manuscript.

## Declaration of Interests

D.B.J. has served on advisory boards or as a consultant for BMS, Catalyst Biopharma, Iovance, Jansen, Mallinckrodt, Merck, Mosaic ImmunoEngineering, Novartis, Oncosec, Pfizer, Targovax, and Teiko, and has received research funding from BMS and Incyte.

## Notes

**Financial Support:** Research was supported by the following funding resources: R01 NS118580 (R.A.I, A.A.B), K00 CA212447 (T.B.), T32 CA009592 (T.B.), the Michael David Greene Brain Cancer Fund (R.A.I.), the Southeastern Brain Tumor Foundation (R.A.I.), a gift from Daniel F. Hewins (R.A.I, A.A.B), and the Vanderbilt-Ingram Cancer Center (VICC, P30 CA68485). The Translational Pathology Shared Resource is supported by NCI/NIH Cancer Center Support Grant P30CA068485 and the Shared Instrumentation Grants S10 OD023475-01A1, S10 OD016355, and IS1BX003154.

**Conflict of Interest Disclosure:** DBJ has served on advisory boards or as a consultant for BMS, Catalyst Biopharma, Iovance, Jansen, Mallinckrodt, Merck, Mosaic ImmunoEngineering, Novartis, Oncosec, Pfizer, Targovax, and Teiko, and has received research funding from BMS and Incyte.

### Competing Interest Statement

DBJ has served on advisory boards or as a consultant for BMS, Catalyst Biopharma, Iovance, Jansen, Mallinckrodt, Merck, Mosaic ImmunoEngineering, Novartis, Oncosec, Pfizer, Targovax, and Teiko, and has received research funding from BMS and Incyte.

### Funding Statement

Research was supported by the following funding resources: R01 NS118580 (R.A.I, A.A.B), K00 CA212447 (T.B.), T32 CA009592 (T.B.), the Michael David Greene Brain Cancer Fund (R.A.I.), the Southeastern Brain Tumor Foundation (R.A.I.), a gift from Daniel F. Hewins (R.A.I, A.A.B), and the Vanderbilt-Ingram Cancer Center (VICC, P30 CA68485). The Translational Pathology Shared Resource is supported by NCI/NIH Cancer Center Support Grant P30CA068485 and the Shared Instrumentation Grants S10 OD023475-01A1, S10 OD016355, and IS1BX003154.

### Author Declarations

IRB of Vanderbilt University Medical Center gave ethical approval for this work (protocol 181970). Statement regarding non-clinical trial status: We confirmed with a treating neurologist in this case that the treatment received by the individual whose samples are studied was part of routine clinical care and not a clinical trial, as the patient was previously diagnosed with Lynch syndrome (a mismatch repair deficiency). The use of pembrolizumab is recommended for treatment of tumors with high mutational burden due to mismatch repair deficiency and is currently considered standard of care for these tumors. Additionally, though the intervention and outcome are detailed in the manuscript, the focus of the manuscript is on reporting changes observed in the immune microenvironment at different points in the clinical trajectory - a retrospective analysis performed after clinical care was complete.

