## Supplementary Figures for "Pembrolizumab alters the tumor immune landscape in a patient with dMMR glioblastoma"

Supplementary Figure 1

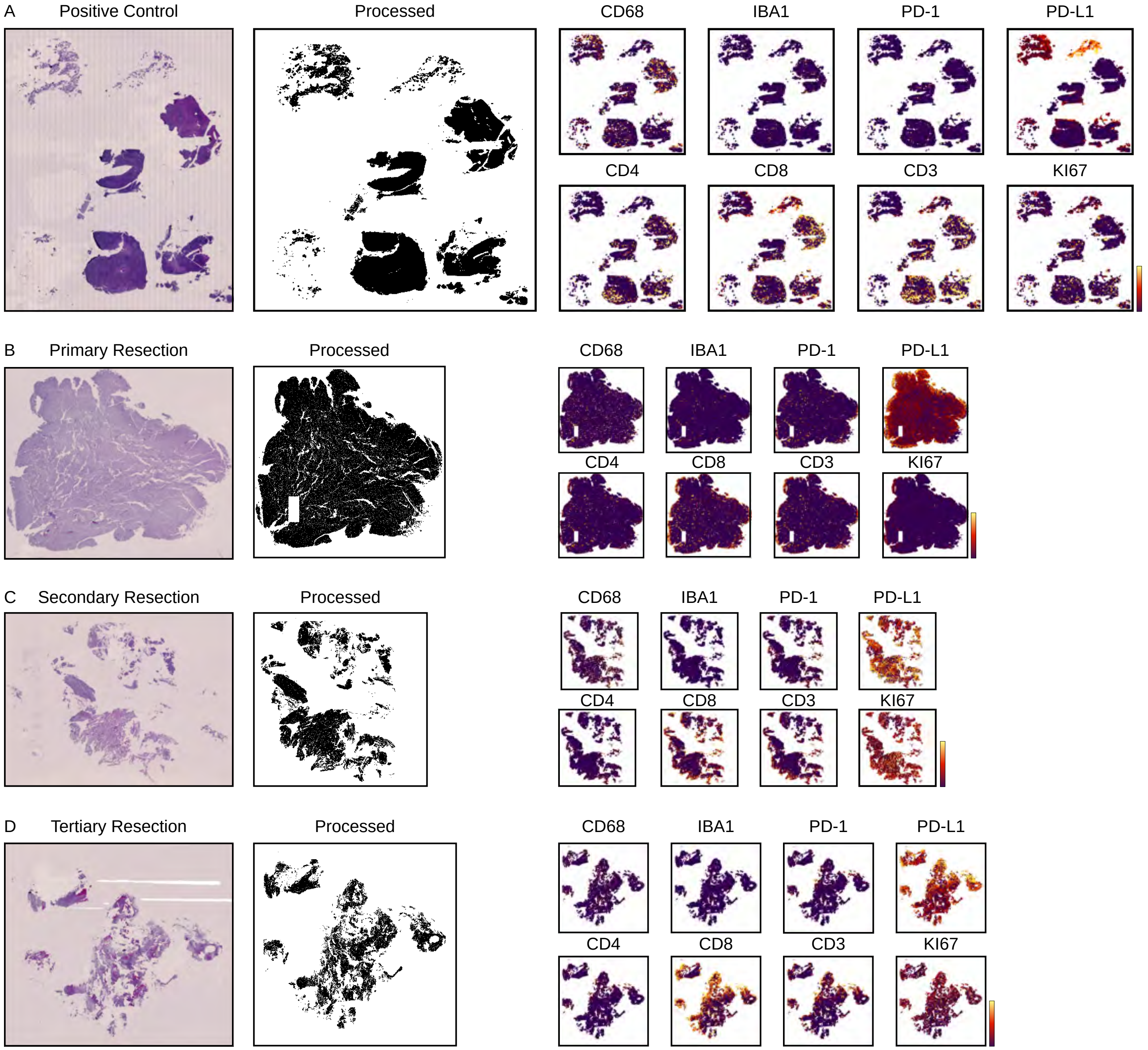

Supplementary Figure 2

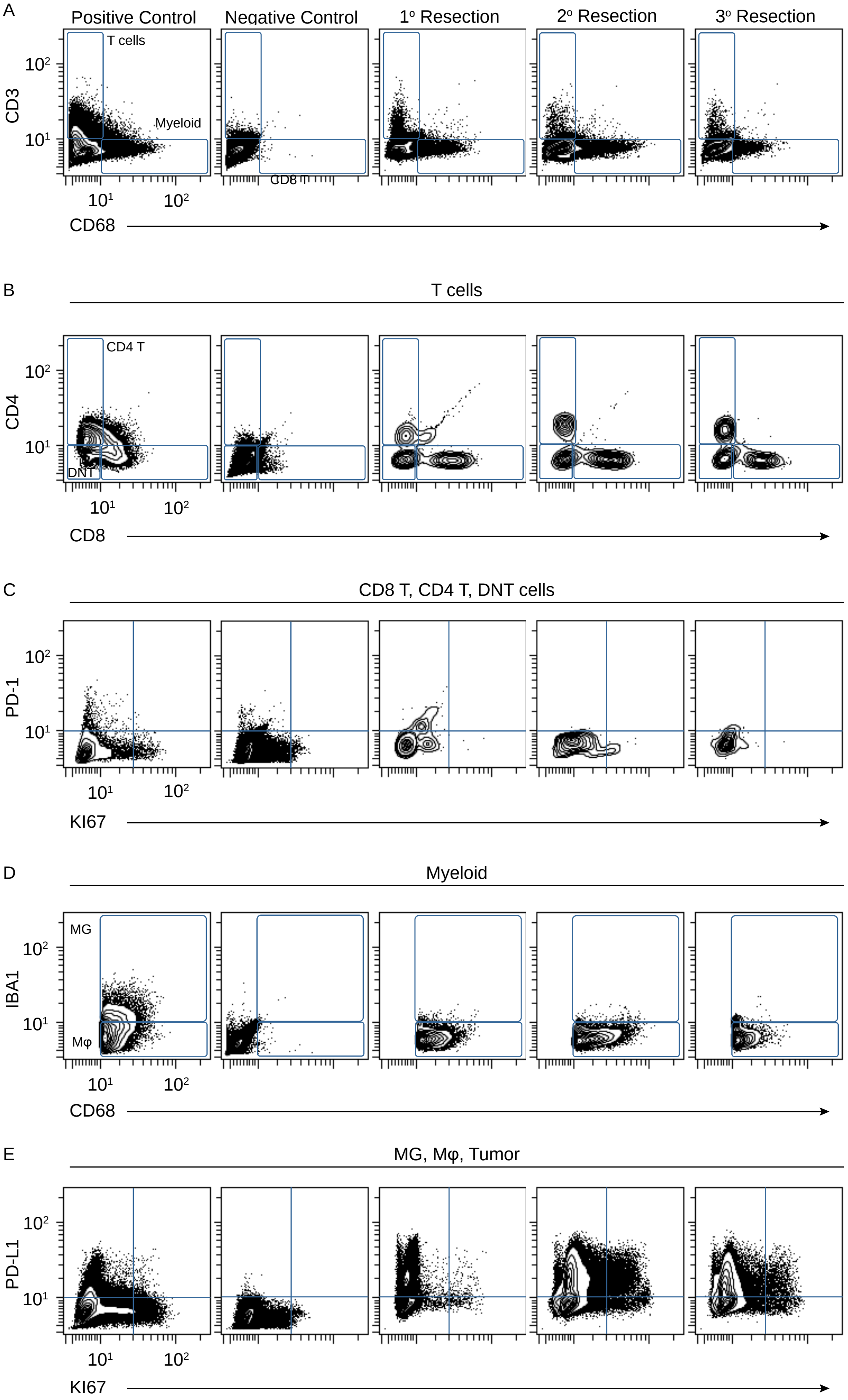

Supplementary Figure 3

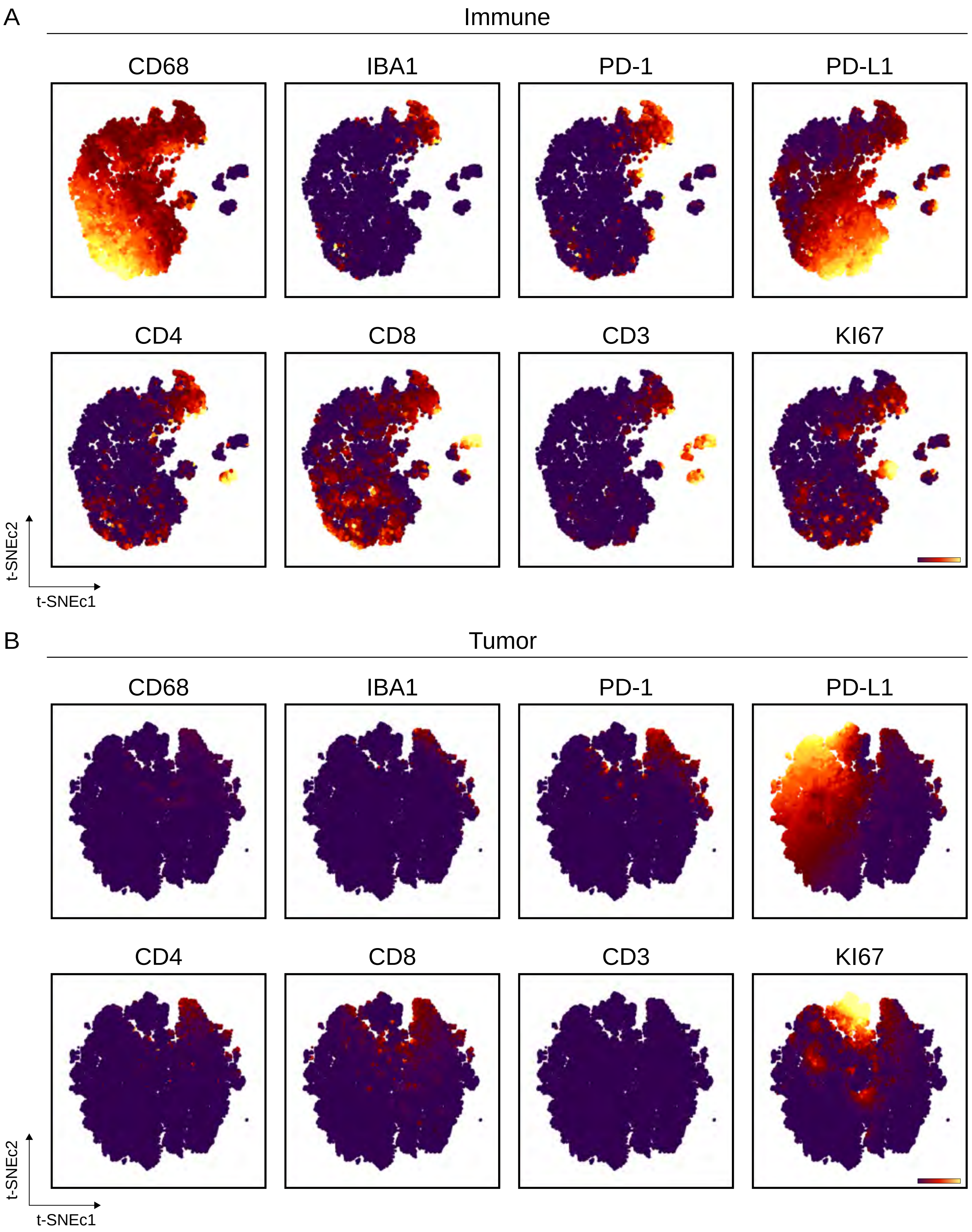

Supplementary Figure 4

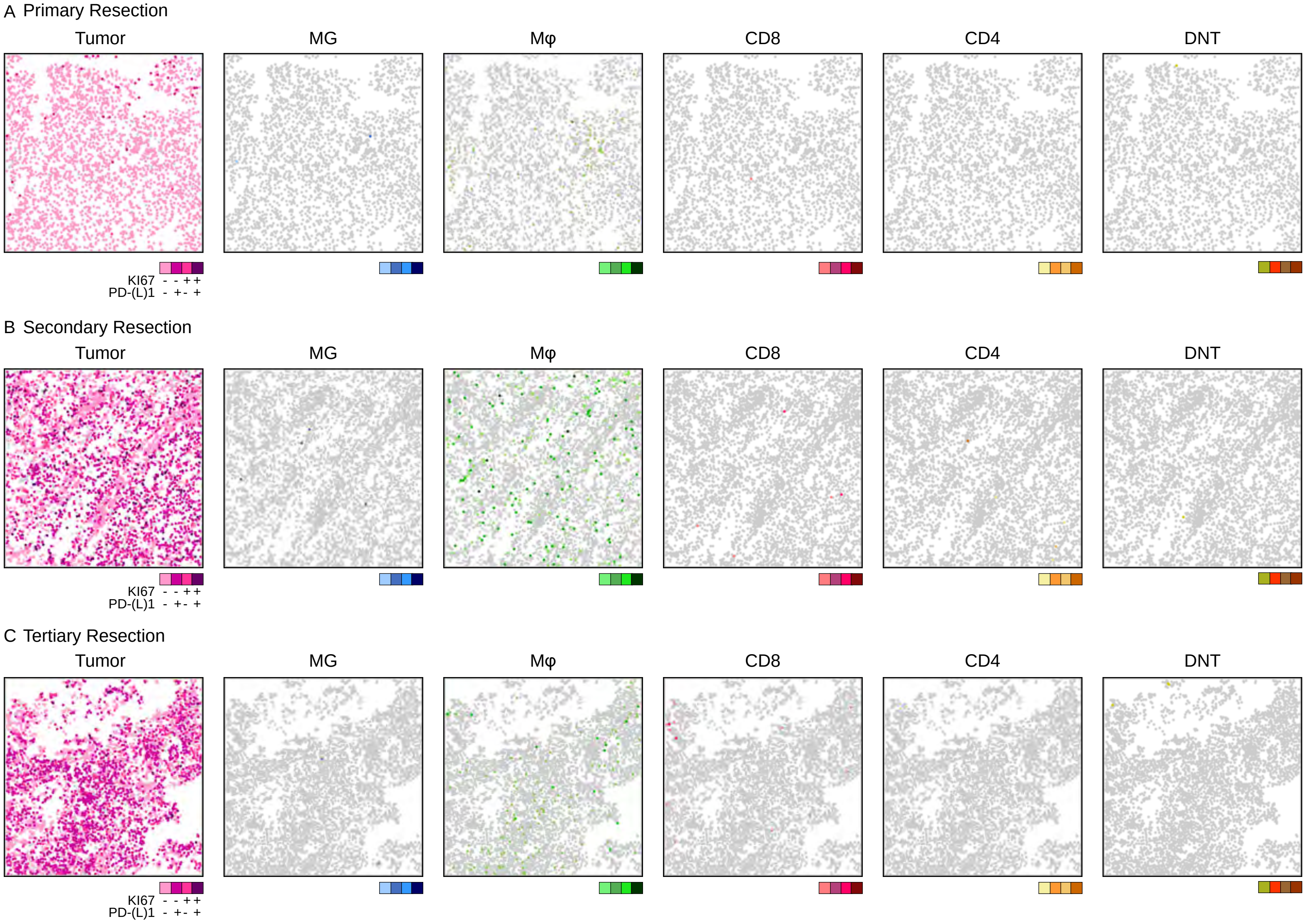

Supplementary Figure 5

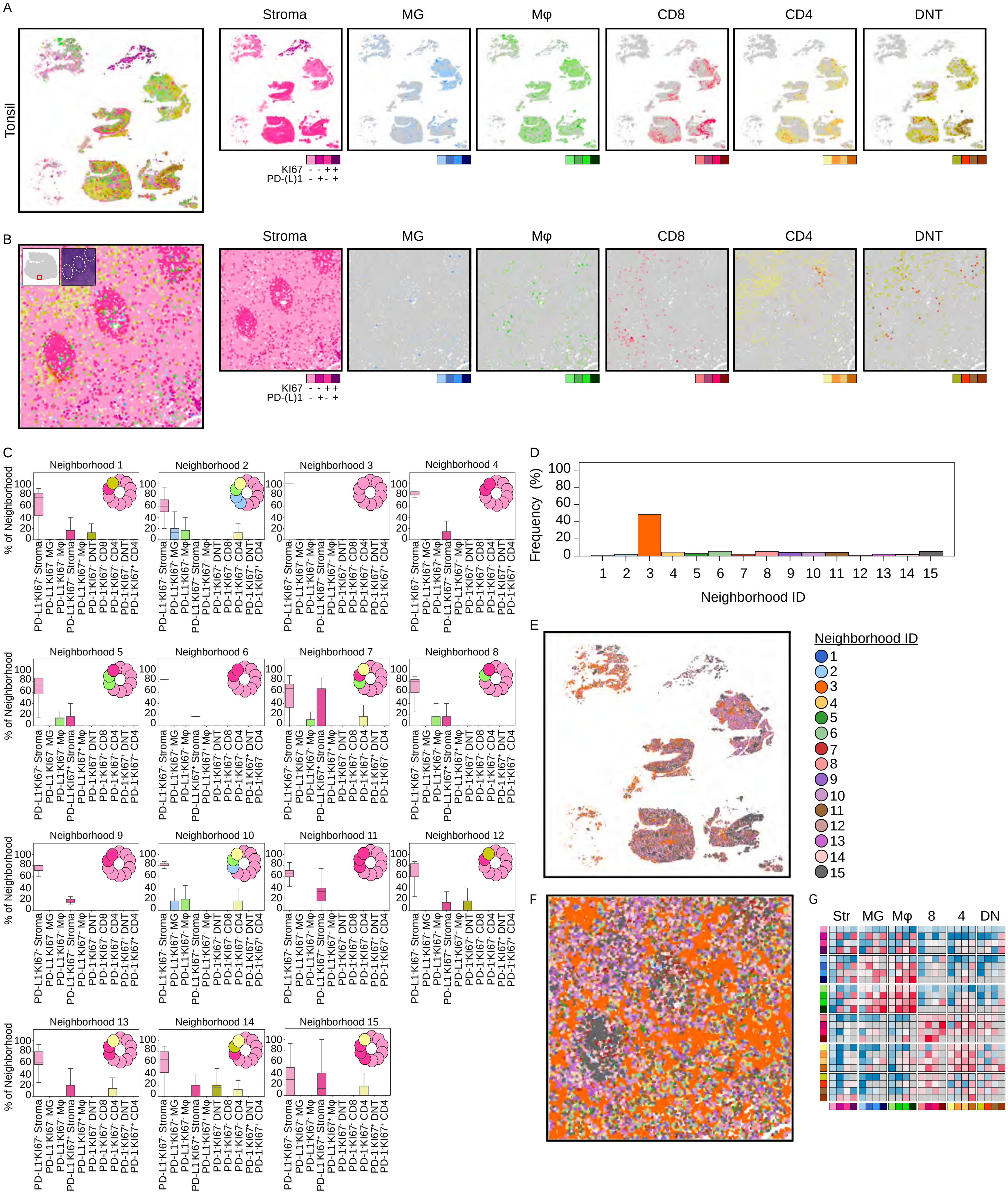

Supplementary Figure 6

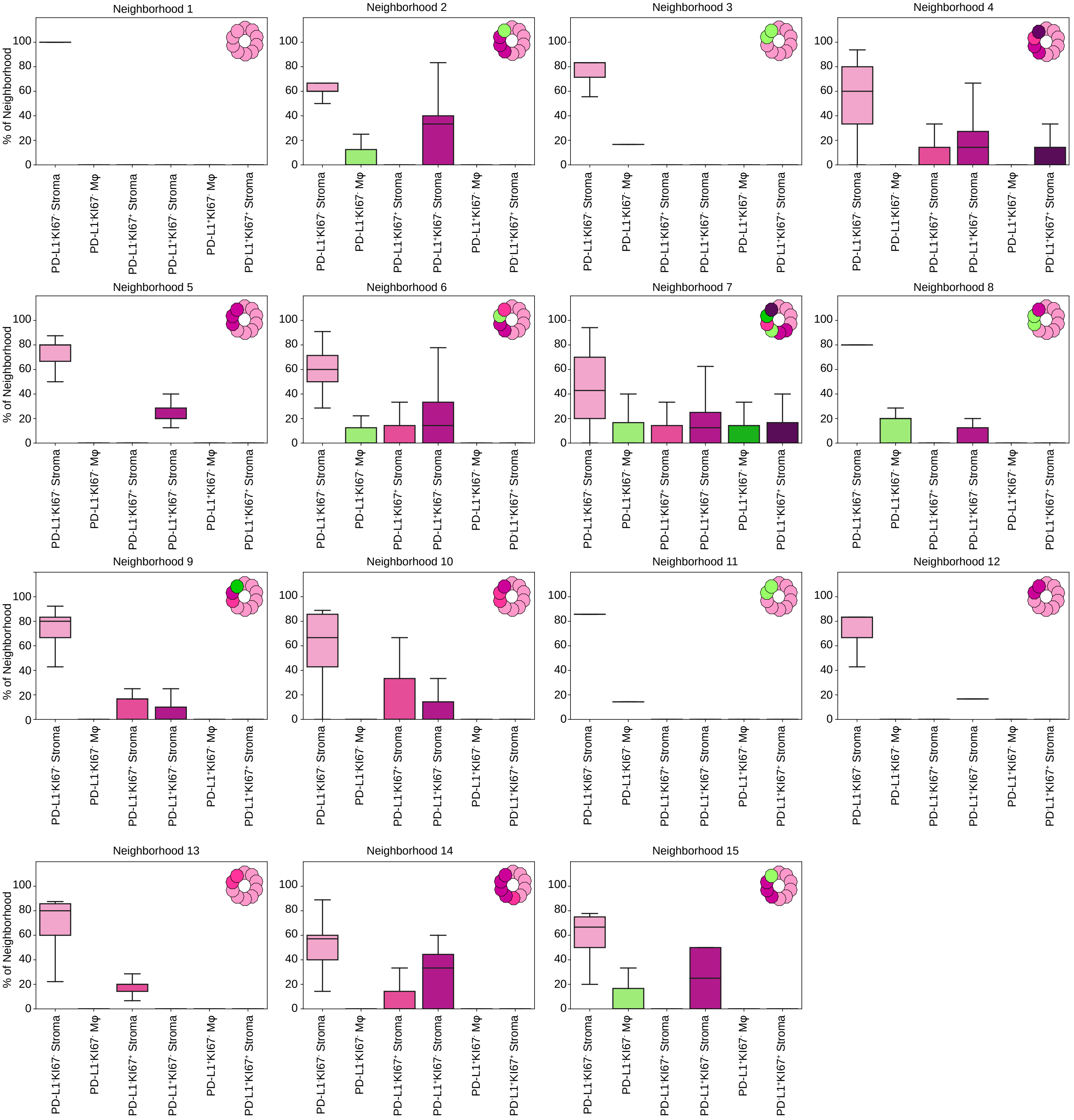

Supplementary Figure 7

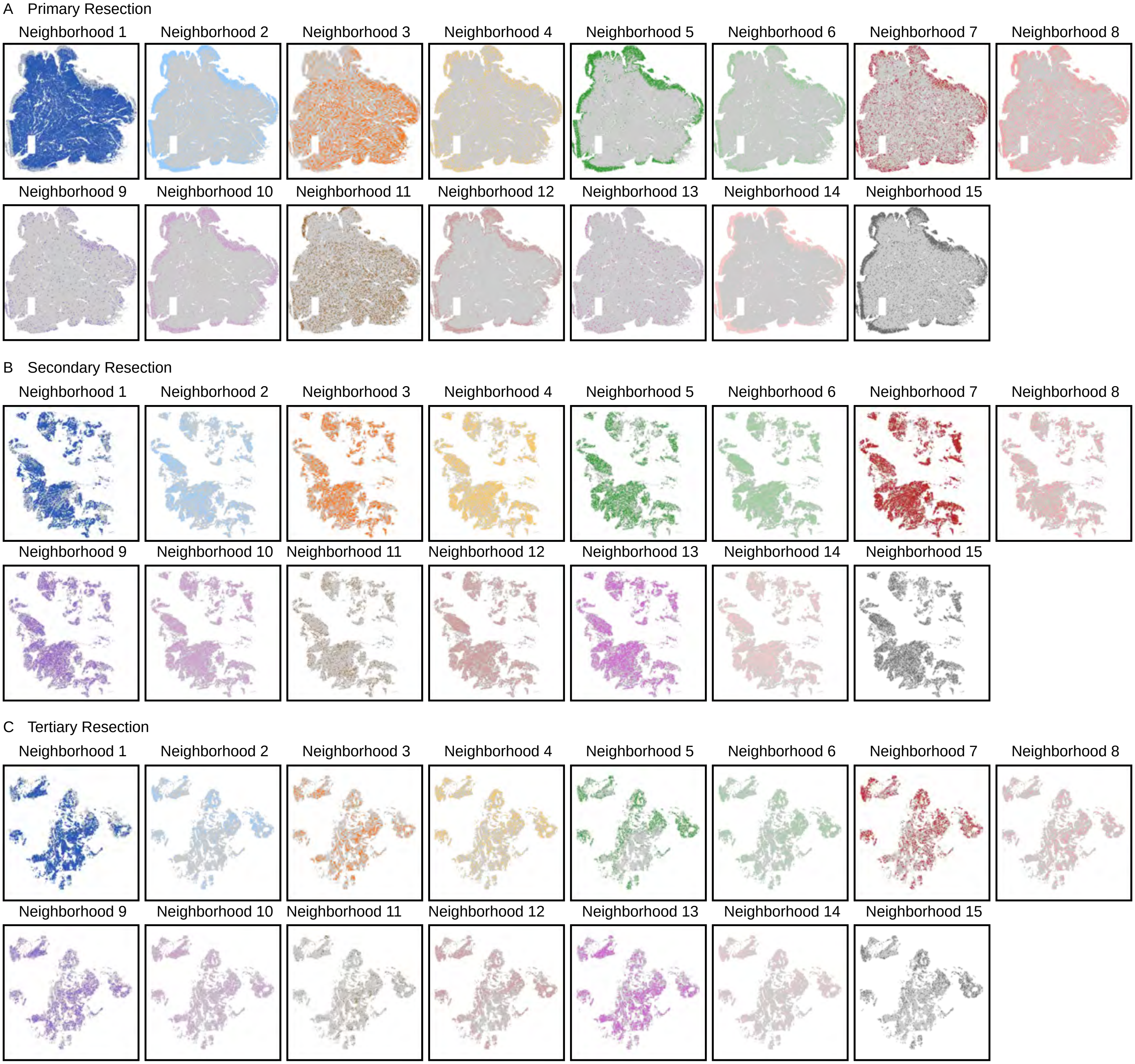

Supplementary Figure 8

A

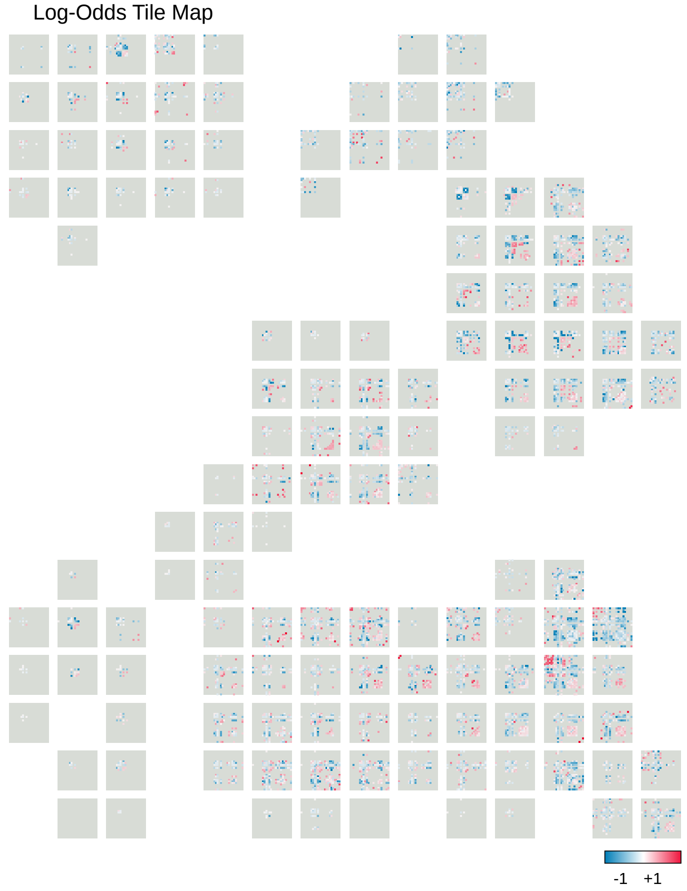


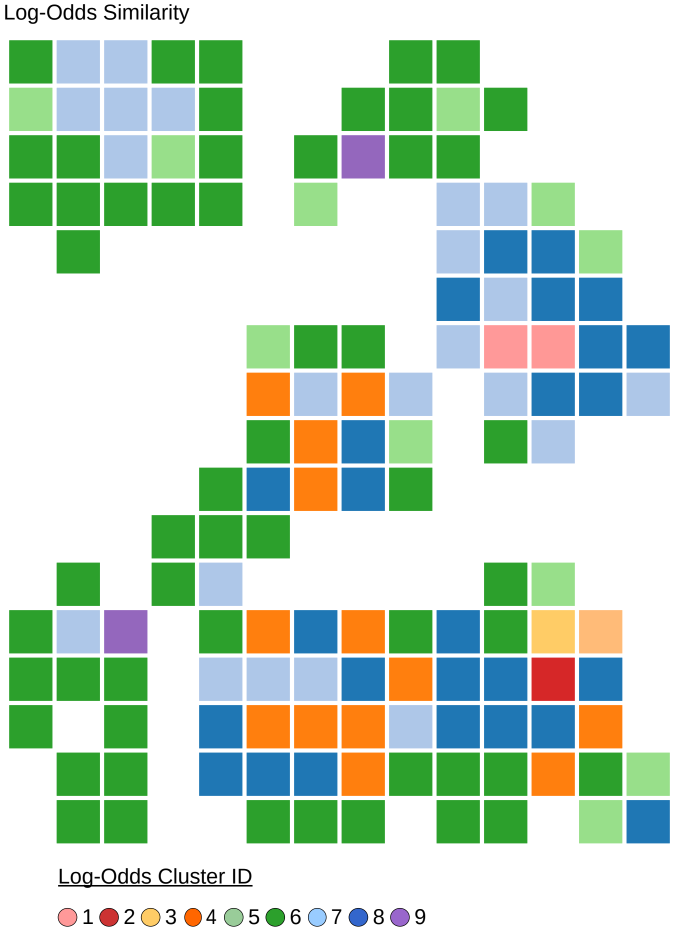

Supplementary Figure 9

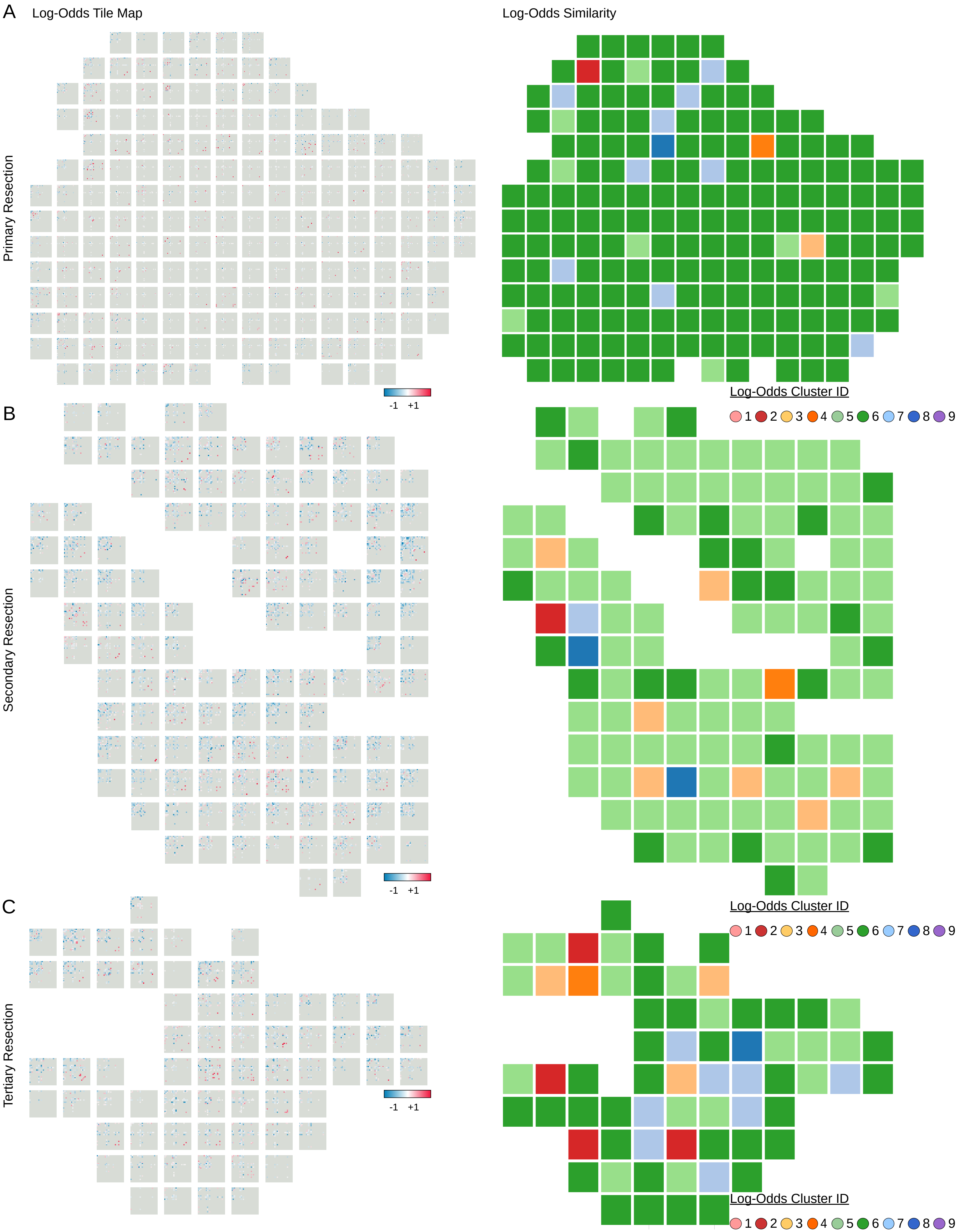
